# Antigenic evidence of lymphatic filariasis transmission in infection persistent endemic districts of Central Nepal during post mass drug administration

**DOI:** 10.1101/2022.11.22.22282615

**Authors:** Pramod Kumar Mehta, Mahendra Maharjan

## Abstract

**Background:** In Nepal, out of 75, 61 districts were endemic with lymphatic filariasis (LF) and completed 6 to 11 rounds of mass drug administration (MDA) with diethylcarbamazine (DEC) and albendazole from 2007 to 2017 in almost all endemic districts of Central Nepal with the aim of eliminating lymphatic filariasis by 2020 but due to fail in transmission assessment Survey (TAS) in some Terai districts of Nepal, aim of elimination was extended to 2030. Antigenemia prevalence have been consistently <2% in all sentinel and check spot sites since 2017 and transmission assessment survey passed in 2018 but in some foci there was low level persistent of infection of *W. bancrofti* was seen in 4 endemic districts of central Nepal so present study has been carried out with the aim of assessing antigenic prevalence in children borne after MDA program to understand evidence of new infection of LF in selected districts

**Methodology and principal findings:** Antigenemia survey was carried out in communities children whose borne after MDA program from selected districts of central Nepal. Two study districts had significantly improved infection to the prior study but two other districts had drastic change. Few hydrocele cases were found but no any children found antigen positive with hydrocele cases.

**Conclusions:** These results indicates that *W. bancrofti* transmission was near to break point in one hilly district (Lalitpur) and one Terai district (Bara) while other two districts from Terai (Mahottari) and hills (Dhading) may requires further intervention. Targeted testing and treatment along with comparative study of children with adult should be required similarly number of MDA rounds should be increased.

**Author summary:** Lymphatic filariasis is neglected tropical disease which cause the disability to human beings which is caused by filarial nematode and transmitted by mosquitoes. Nepal government was completed 6-11 rounds of antifilarial medicine with DEC and albendazole between 2007 to 2017 in all endemic districts of central Nepal. TAS report showed low level of infection in all study sites. The present study reports assessed antigenic prevalence of 4 infection persistent endemic districts of central Nepal with high signals than prior study in 2 areas while other 2 areas showed in elimination phase so further investigation along with number of MDA rounds should be required to interrupt transmission in high antigenic prevalence districts. Our results suggest that further xenomonitoring and microfilaremia study should be required for informed decision, targeted test and treatment should required.

## Introduction

Lymphatic filariasis (LF) is the second most common vector-borne parasitic disease after malaria, and is found in more than 80 tropical and subtropical countries [1].The disease is caused by three filarial nematodes: *Brugia malayi, Brugia timori*, and most commonly, *Wuchereria bancrofti* [2]. All these parasites are found in different physiological races based on the microfilaria in the peripheral blood, the species are further classified into the periodic (presence of microfilaria during either day or night in humans) and sub periodic (presence of microfilaria all the time with a peak either during day or night), the parasite with a given physiological race is transmitted by different species of mosquitoes belonging to four principal genera—*Anopheles, Culex, Aedes and Mansonia* [3]. in different regions of the tropics, although more than 50% of the LF infections all over the world are transmitted by a single vector species, *Culex quinquefasciatus* [4].

Lymphatic filariasis caused by filarial parasites remains an important public health problem in a 83 countries and globally, 1.3 billion people are at risk of infection with about 120 million people are affected [5].. This tropical disease manifesting as different health states causes long-term suffering and morbidity as well as high social and economic burden to individuals and communities [6-9].). Even though the disease is not fatal, it is ranked as the second leading cause of disability [10, 11]. and imposes heavy burden on the health care infrastructure in endemic areas [12].. Three Asian countries, India, Indonesia, and Bangladesh as well as a African country Nigeria contribute about 70% of the infection Worldwide [13].

Lymphatic filariasis was identified as one of the six parasitic diseases which could be potentially eradicated [13]. Preventive chemotherapy using diethylcarbamazine (DEC) and albendazole is recommended for interrupting transmission of LF and repeated rounds of treatment could reduce the parasite load in the community with the aim of elimination of Lf by 2030. In mass drug administration, all eligible people in all endemic areas are given a single dose of two drugs (DEC and Albendazole) together once a year for at least 5 years. By the end of 2011, 53 of 73 endemic countries were implemented mass drug administration, and more than 3.9 billion treatments had been delivered to 952 million people [5].

Nepal government had formulated national task force (2003-2020AD) and launched global programme to eliminate lymphatic filariasis in 2003 with the support of WHO (DHS 2017). Main focus of the programme was to interrupt transmission of microfilaremia in the community by mass drug administration (MDA) in 2020 but due to some reason elimination was not possible in 2020 so Nepal government taken new target to eliminate in 2030. Nepal is administratively divided into 75 districts, of which 61 districts (23 districts are in the Terai region and 38 are in hills with mountain) are potentially endemic for lymphatic filariasis and remaining 14 districts are in mountainous region and unlikely to be endemic since transmission does not normally occur above 2,000 meters [14, 15]. (DHS 2017).

In Nepal MDA programme initiated in 2003 had covered six rounds of MDA in 51 districts in 2017 while remaining districts have targeted to complete in coming year i.e. 2018 (DHS 2017). Post MDA surveillance report showed in 2016 in 31 districts Ag prevalence was < 1% so results of TAS indicated that though the 31 districts were qualified for stopping MDA (EDCD unpublished report). Though there are foci of persistent infection with residual microfilaria carriers and transmission evidenced by antigen positive children. These districts are under post MDA surveillance. Monitoring is required to be continued for five more years with swo more TAS with a gap of two years. The district will be declared free from transmission and qualified for certification for elimination only when the subsequent TAS showed absence of transmission. Therefore it is important to monitor the situation, so as to ensure that there is no resurgence of infection. In this contest it is important to understand the status of persistent infection areas in the risk of transmission and possible resurgence. This study will thus carry out in 4 most endemic districts of central Nepal; Where MDA surveillance to assess the risk of transmission is under post MDA and recommended appropriate method of monitoring the situation in infection persistence districts. A present study aimed to cover four districts and around the house of antigen (Ag) positive cases will be proposed to see all the individuals to assess the prevalence of antigen in the selected age class.

## Methods

### Ethical review and consent to participate

Field surveys were conducted as per the study protocols approved by the Nepal Health Research Council (NHRC/Reg.no. 629/2018). Informed consent was obtained from all the individuals screened for antigenemia. Information on the objective of the study and the expected benefits to the individual, and the community were provided.

### Study sites

Area with persistent infection can be considered as “hot spot” The risk of infection in such “hot spot” is to be investigated in children whose borne after MDA to suggest new infection status of Lf. Out of 19 districts of central Nepal, 9 districts are in Teri, 7 districts are in hills and 3 districts are in mountain region in which 17 districts were endemic with Lf while only 2 districts of mountain were non endemic with LF in central Nepal. Based on TAS 2017 report (Unpublished data of MOHP) 4 districts of central Nepal, 2 from hilly (Lalitpur and Dhading) and 2 from Terai region (Bara and Mahottari) were selected for study while sentinel sites of TAS of these districts were purposively selected for study.

Rural area of Lalitpur district (27^0^40’N, 85^0^19’E) of Kathmandu valley were selected for study Based on 2011 census data the district had a total population of 513,200 people (CBC 2011). The district had elevated tract of land on the south side of the Bagmati River which separated from Kathmandu. The district has two main rainy season, a long rainy season between April and September while shorter less instance from December to February with annual rainfall has 1343mm. Eight rounds of annual MDA were completed in between 2010 and 2017 (DHS 2017).Lalitpur district had six municipalities including three rural municipalities and one metropolitan city. Using purposive sampling, two villages Bugmati of Lalitpur metropolitan city and Dhukuchhap of Godawari municipality were selected for study. The present study was conducted in May 2019, about after three years of 8 rounds of MDA based on a combination of DEC and albendazole delivered in September 2017 while this study was also conducted in Tripurasundari rural municipality located within the Dhading district of hilly region of central Nepal. This rural municipality has a total population of 22960 and area is 271.23 square KM based on 2011 Nepal census (CBC, 2011) while MDA was started 2007 and six rounds completed in 2013

Bara district (27^0^ 01’ 60.00” N, 85^0^ 00’ 0.00” E) lies in Terai region of central Nepal and Kalaiya as its headquarters covers an area of 1.190km^2^ while it has a population of 687,708 (CBC 2011) where this study was conducted in Ammadar and Khairawa village of Jeetpur sub-metropolitan city in Bara district while this study was conducted in December 2021 about after eight years of 6 rounds of MDA based on a combination of DEC and albendazole delivered in September 2013.

Mahottari district lies in Terai region of central Nepal where Matihani Municipality (26°36’50.0”N 85°50’50.1”E) was sentinel sites for TAS where this study was completed in April 2022 about after nine years of 6 rounds of MDA based on a combination of DEC and albendazole delivered in September 2013 while Jaleshwar as its headquarters, covers an area of 29.02 sq. km and has a population of 31,026 (CBC 2011).

No any treatment of filariasis was provided in this interval in 3 districts but one round of MDA with DEC and Albendazole was provided in one district (Bara) in 2020. A study map showed in fig 1.

**Fig 1.**
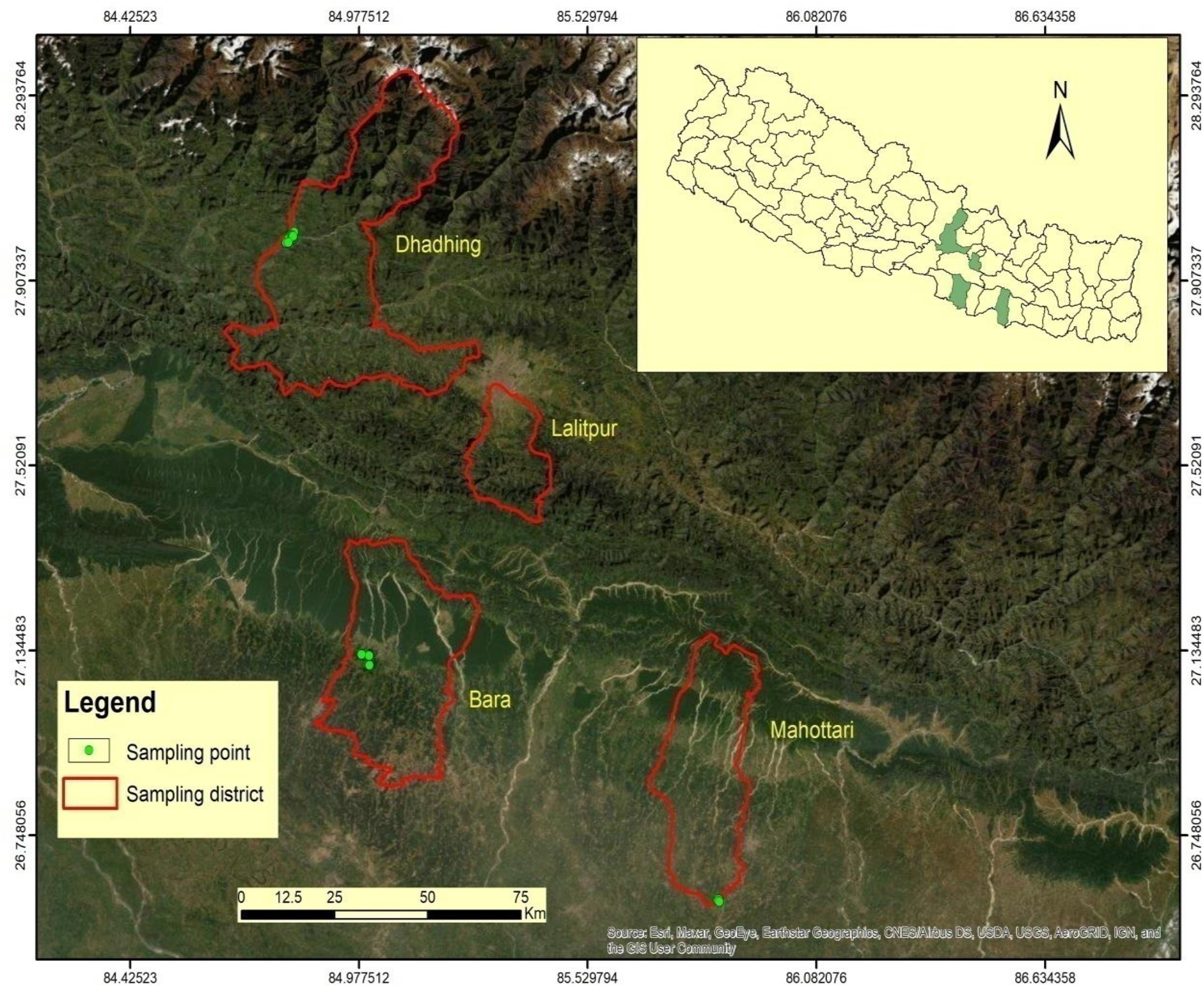
The map shows 4 filarial endemic districts with approximate location of sampling points with green circles that were surveyed in 2019-2022.

### Study Population and Sampling

Participants were selected from list of selected households which was obtained from respective administrative officer. A simple random sample of 724 households was selected (186 in Lalitpur, 174 in Dhading, 141 in Bara and 223 in Mahottari) with a target sample size of 692 individuals. All individuals 5-15 years of age groups of children whose borne after MDA to participate and tested for filarial antigenemia using ICT (FTS Alere, Scarborough, USA) at day time. A standard questionnaire survey was done during a time of testing to collect data on demographics and MDA compliance history. All participants involved in the survey provided written consent before participation.

### Antigenemia testing for human subjects

Antigenemia testing was performed using Filarial Test Strip (Alere, Scorborough ME) by fingers prick method. In a plastic micropipette, 75 μL of blood was directly placed onto the Filarial Test Strip(FTS) sample application pad and a single operator was read the FTS at 10 minutes. Visual Filariasis Test Strip (vFTS) results was scored semi quantitatively. Each of the strip was labeled with patient ID, date and result scored.

### Data analysis

Data were entered in Excel spreadsheets (Microsoft excel 2007) and subsequently analyzed with Minitab 17 version 19.2.0.Blood test results for CFA (Circulating Filarial Antigen) and MF, the presence of hydrocele or elephantiasis, demographic characteristics and MDA compliance were compared by using the Chi-square test and Fisher’s exact test while p-value ≤ 0.05 were considered statistically significant. The lower and upper limits of the 95% CI for the prevalence of CFA were calculated.

## Results

### Community survey results

Four districts from 2 regions (Hilly and Terai) were surveyed for *W. bancrofti* infection parameter between 21 May 2019 to 17 April 2022. Survey periods for sample were in Lalitpur (May, June 2019), Dhading, (Nov, Dec, 2019), Bara (Dec, Jan 2022) and Mahottari (April, May 2022). A total of 791 people from the community were surveyed (age 5-15 years). Average mean age was 9.2 years, 44% males. Out of 4 districts Bara district MDA was stopped after maximum MDA rounds of 11 while in Dhading and Mahottari it was stopped at 6 MDA rounds. In both hilly and Terai districts median coverage and IQR were significantly associated (Table 1).

**Table 1:** Reported median treatment coverage during DEC and albendazole MDA intervention in selected districts of Nepal.

|  | Districts | Number of MDA rounds | Median coverage of MDA | IQR | P-value |
| --- | --- | --- | --- | --- | --- |
| Hilly | Lalitpur | 8 | 61.4 | 19.8 | 0.002* |
|  | Dhading | 6 | 80.8 | 5.5 |  |
| Terai | Bara | 11 | 82.6 | 4.1 | 0.044* |
|  | Mahottari | 6 | 87.3 | 7.6 |  |
| MDA, Mass drug administration, IQR, Inter quartile range, DEC, Diethylecarbamazine, P values with “*” symbol were statistically significant. P values shown in results column are for differences compared to median coverage of MDA |  |  |  |  |  |

In present survey out of eligible population around 70% were sampled from both sexes covering age group of 5 to 15 years. Maximum eligible population was covered from Dhading and Bara districts for antigenic survey (Table 2).

**Table 2.** Demographic characteristics of eligible sampled and missed population of lymphatic filariasis in selected districts of central Nepal during post MDA.

| Demographic characteristics |  | Eligible population | Sampled population (%) | Missed population (%) |
| --- | --- | --- | --- | --- |
| Gender | Male | 615 | 432 (70.24) | 183 (29.76) |
|  | Female | 502 | 359 (71.51) | 143 (28.49) |
| Age group<br>(years) | 5-9 | 652 | 436 (66.87) | 212 (32.52) |
|  | 10-15 | 465 | 355 (76.34) | 110 (23.66) |
| Hilly | Lalitpur | 375 | 176 (46.93) | 199 (53.07) |
|  | Dhading | 208 | 202 (97.12) | 6 (2.89) |
| Terai | Bara | 227 | 211 (92.95) | 16 (7.05) |
|  | Mahottari | 307 | 202 (65.80) | 105 (34.20) |

Upper confidence limit of CFA prevalence was greater than 2% in all study districts indicating risk of resurgence of Lf transmission. Comparatively Dhading district of hilly region and Mahottari district of Terai region have antigenic prevalence significantly high (p= <0.05) (Table 3).

**Table 3.** *W. bancrofti* antigenemia prevalence of lymphatic filariasis in districts of central Nepal during post MDA.

|  | Districts | CFA positive/Total (% 95% CI) | Fisher’s exact test (P-value) | Prevalence (%) |
| --- | --- | --- | --- | --- |
| Hilly | Lalitpur | 1/176 (0.0-3.1) | ≤0.001* | 0.6 |
|  | Dhading | 20/202 (6.2-14.9) |  | 9.9 |
| Terai | Bara | 2/211 (0.1-3.4) | ≤0.001* | 1.0 |
|  | Mahottari | 26/202 (8.6-18.3) |  | 12.9 |
| CFA: Circulating Filarial Antigen. *W. bancrofti* infections were detected by antigen (CFA) testing. % of antigen prevalence were significantly lower in Lalitpur and Bara districts*. |  |  |  |  |

No significance difference was found in different age and sex group in Dhading and Mahottari but significance difference was found in Lalitpur and Bara (P=≤0.002). Few hydrocele cases (8 of 791) were identified during survey but no any individuals found with elephantiasis. None of the individuals with hydrocele were found to have CFA (Table 4)

**Table 4.** Summary of circulating filarial antigen and clinical manifestation of lymphatic filariasis infection in study areas.

| Characterstics |  | No. of examined | Prevalence | CFA 95%CI | P-value | Hydrocele % |
| --- | --- | --- | --- | --- | --- | --- |
| Lalitpur |  |  |  |  |  |  |
| Gender | Male | 108 | 0.9 | 0.02-5.1 | ≤0.001* | 1.9 |
|  | Female | 68 | 0.0 | 0.0-4.3 |  | - |
| Age group<br>(Years) | 6-7 | 74 | 1.4 | 0.03-7.30 | ≤0.001* | 0.0 |
|  | 8-9 | 102 | 0.0 | 0.0-4.3 |  | 2.0 |
| Dhading |  |  |  |  |  |  |
| Gender | Male | 105 | 11.4 | 5.3-17.5 | 0.493 | 1.9 |
|  | Female | 97 | 8.2 | 2.8-13.7 |  | - |
| Age group<br>(years) | 5-9 | 108 | 9.3 | 3.8-14.8 | 0.767 | 0.0 |
|  | 10-12 | 94 | 10.6 | 4.5-16.9 |  | 1.9 |
| Bara |  |  |  |  |  |  |
| Gender | Male | 114 | 0.0 | 0.0-2.6 | ≤0.001* | 1.8 |
|  | Female | 97 | 2.1 | 0.25-7.3 |  | - |
| Age groups<br>(years) | 5-9 | 124 | 1.0 | 0.2-5.7 | ≤0.001* | 1.6 |
|  | 10-15 | 87 | 0.0 | 0.0-3.4 |  | 0.0 |
| Mahottari |  |  |  |  |  |  |
| Gender | Male | 105 | 16.2 | 9.7-24.7 | 0.197 | 1.9 |
|  | Female | 97 | 9.3 | 4.3-16.8 |  | - |
| Age groups<br>(years) | 5-9 | 92 | 10.9 | 5.3-19.1 | 0.494 | 0.0 |
|  | 10-15 | 110 | 14.6 | 8.6-25.5 |  | 1.8 |
| CI, Confidence interval, P values with”*” symbols were statistically significant. P values are based on Chi-squire analysis and Fisher’s exact test. |  |  |  |  |  |  |

### Negative association between filarial infection prevalence and MDA coverage

Out of 2 districts from hilly region, median coverage of MDA was significantly more in Dhading (p=0.002), similarly out of 2 districts from Terai region, median coverage of MDA was significantly more in Mahottari (p=0.044) but CFA prevalence was significantly more in Mahottari and Dhading (p=≤0.002) so CFA prevalence was not associated with MDA coverage. Filarial infection prevalence was directly associated with number and recent MDA rounds

## Discussion

WHO launched the global program to eliminate lymphatic filariasis in 2000 following the resolution of World Health Assembly 2007 and targeted for elimination globally by 2020 but due to some reason target for elimination is not fulfill so GPELF set the new NTD road map 2021-2030 to eliminate Lf. Preventive chemotherapy using mass annual single dose DEC and albendazole is the main transmission control strategy [16]. in preventing new infections thereby achieving elimination. All the three filarial parasites viz., *Wuchereria bancrofti, Brugia malayi* and *Brugia timori* with their physiological races of periodic and sub periodic are known to respond well to the drugs used in the programme. About 1380 million people were at risk of infection in 72 countries which were earlier known to be endemic [17], ten countries were classified as non-endemic with no evidence of indigenous transmission. Fifty eight countries have successfully interrupted the transmission and are under post-MDA surveillance [17]. When the programme was launched, guidelines for programme planning and implementation [18] and monitoring and evaluation, particularly on the decision making were available. However, tools and protocols for monitoring and evaluation [19] were not operationally feasible with highly conservative levels of threshold. As the programmes advanced with up scaling, inputs from the programme and researchers [20-26].were used and in 2011 a revised protocol viz., Transmission Assessment Survey was developed and recommended for monitoring and evaluation of LF elimination programme [27].. The immunochromatographic test based tools recommended for assessment and verification of absence of transmission during post MDA period for detecting filarial antigen of *W. bancrofti*.

The major operational issue identified as a limiting factor for effective implementation of MDA was compliance. Achieving requires level of coverage and consumption of drug is crucial for the success of the programme as it is known to influence the impact of MDA [28] which is shown to be directly related to compliance from a model based study [29]. Minimum effective coverage of the total population was estimated to be 65% [30]. Though five rounds of MDA have been recommended to the minimum target, a systematic review on the mass chemotherapy options to control LF [31] and other studies have shown that the required number of rounds of MDA would depend on the initial prevalence of infection, initial intensity of transmission, the efficacy of drugs, the combination of parasites and vectors and density of vectors [32-36].

Subramanian et al (2012) have reported existence of foci of persistent infection and transmission which are defined as presence of infection (Mf and or antigenemia) above 1% level or prevalence of antigenemia children in the age class of 2-8 years [26]. These areas require focus during post MDA surveillance so as to detect the signals for possible resurgence of infection. The results of long-term observations on the status of such areas are yet to be known. The present study is an attempt to understand the status of transmission in the communities with persistent infection (residual Ag positive children). Therefore use of antigenemia as indicator particularly community based, may lead to underestimation of the impact of MDA, particularly after a few rounds of MDA as observed by Schuetz et al (2000) Tisch et al (2008) and Simonson et al (2010) [37-39].

The present study in the community of 2 hilly districts (Lalitpur and Dhading) and 2 Terai districts (Bara and Mahottari) with antigenemia prevalence ≥ 2% in Dhading and Mahottari show evidence of recent infection but only one of the children below 15 years was found positive for antigenemia in Lalitpur which was migrated from Banke endemic district of Nepal that indicating the absence of recent transmission. Antigenemia prevalence was below the critical level in Bara (≤2%) so there is no risk of resurgence of infection but area is receptive so post MDA surveillance is required. The strength of our study was the method used to conduct CFA prevalence surveys was well supervised and standardized. However there are some limitation because the use of already collected data. We were not able to verify the accurately MDA uptake of risk population. Median coverage of MDA was less in Lalitpur and Bara districts in comparison with Dhading and Mahottari districts but numbers of MDA rounds were more in Lalitpur and Bara districts so our results indicating LF prevalence directly associated with number of MDA rounds.

Therefore this area demands to strengthen of monitoring, testing and treating for high risk populations can be a potential option. Vector control could be yet another solution to ensure that recrudescence will not occur from the residual infection. However, there could be limitation in carrying out post MDA surveillance activities in situation which are constrained with manpower. Integrating post MDA surveillance activities with those of other Neglected Tropical Diseases (NTDs) control programme or integrating LF surveillance activity with population based surveys could minimize the need for long-term resources for LF specific surveillance [40].

Mass distribution of DEC salt could be an alternative intervention in the transmission hot spots to prevent recrudescence as recommend to hasten the process of LF elimination [41, 42].

## Conclusions

The variability in district level antigenemia within 2019 to 2022 surveys suggest that there remains significant gap in knowledge regarding spatial transmission dynamics at district level. For better understanding further research on transmission dynamics should required for accurately assess the significant residual infection during post-MDA phase that can lead to elimination of Lf in countries.

Analysis of Lf infection indices in children demonstrated substantial decline in antigen prevalence in one hilly and one Terai district(Lalitpur and Bara) in compared with baseline prevalence so there is no risk of resurgence of infection but area is receptive so further monitoring should required during post-MDA situation but in one district from Terai and one from hilly (Mahottari and Dhading) districts antigen prevalence was significantly more in compared with baseline prevalence so there is new infection that leads to further risk of resurgence of infection. We recommended further post TAS to be conducted to make informed decision on whether continued. Targeted testing and treatment should required along with comparative testing of antigen prevalence in children with adult should required.

## Data Availability

All relevant data supporting the conclusions of this article are included within the article

## Abbreviations

LF: Lymphatic filariasis
MDA: Mass drug administration
CFA: Circulating filarial antigens
MF: Microfilaria
Ag: Antigen
ICT: Immunochromatographic test cards
TAS: Transmission assessment survey
WHO: World health organization
CDC: Centers for disease control and prevention
GPELF: Global Programme to Eliminate Lymphatic Filariasis

## Acknowledgements

The authors extend their extremely grateful thanks to the residence of study area who participated in the study. We express deepest thanks to Prof. Dr. Tej Bahadur Thapa, Head of Department, Central Department of Zoology (CDZ), Tribhuvan University (TU), Nepal for his valuable suggestions. The authors wish to thanks all technical volunteers and female community health volunteers for helping in data collection from study area. We would like to thank Mr. Jagan Nath Adhikari of CDZ, TU, Nepal for helping in ArcGIS maps.

## Availability of data and materials

All relevant data supporting the conclusions of this article are included within the article

## Authors’ contribution

PKM conceived, designed, ran the experiments, analyzed the data, and wrote the manuscript. MM helped during writing the manuscript. All authors read and approved the final manuscript

## Consent for publication

Not applicable

## Supporting information

**S1 Fig. Sample collection by finger prick method for antigenemia study in field**

**S2 fig: A positive test read at 10 minutes**

